# Symptom reporting in over 1 million people: community detection of COVID-19

**DOI:** 10.1101/2021.02.10.21251480

**Authors:** Joshua Elliott, Matthew Whitaker, Barbara Bodinier, Steven Riley, Helen Ward, Graham Cooke, Ara Darzi, Marc Chadeau-Hyam, Paul Elliott

## Abstract

Control of the SARS-CoV-2 epidemic requires rapid identification and isolation of infected individuals and their contacts. Community testing in England (Pillar 2) by polymerase chain reaction (PCR) is reserved for those reporting at least one of four ‘classic’ COVID-19 symptoms (loss or change of sense of smell, loss or change of sense of taste, fever, new continuous cough).^1^ Detection of positive cases in the community might be improved by including additional symptoms and their combinations. We used data from the REal-time Assessment of Community Transmission-1 (REACT-1) study to investigate symptom profiles for PCR positivity at different ages. Among rounds 2–7 (June to December 2020), an age-stratified, variable selection approach stably selected chills (all ages), headache (5–17 years), appetite loss (18–54 and 55+ years) and muscle aches (18–54 years) as jointly and positively predictive of PCR positivity together with the classic four symptoms. Between round 7 (November to December 2020) and round 8 (January 2021) when new variant B.1.1.7 predominated, only loss or change of sense of smell (more predictive in round 7) and (borderline) new persistent cough (more predictive in round 8) differed between cases. At any level of PCR testing, triage based on the symptoms identified here would result in more cases detected than the current approach.

## Main

High community transmission and prevalence of severe acute respiratory syndrome coronavirus 2 (SARS-CoV-2) in England during December 2020 and January 2021^2^ led to a third national lockdown starting 6 January 2021. To help control the epidemic, rapid identification and isolation of infected individuals is essential to curtail the transmission of SARS-CoV-2^3^ together with testing and isolation of coronavirus disease 2019 (COVID-19) contacts.^4,5,6^ Polymerase chain reaction (PCR) is the gold standard^7^ for SARS-CoV-2 detection in the general population. The UK government has established a National Health Service Test and Trace system for testing symptomatic individuals in the community (Pillar 2) based on reporting of at least one of the following four ‘classic’ COVID-19 symptoms: fever, new continuous cough, loss or change in sense of smell or taste.^1^ We hypothesised that detection of cases in the community might be improved by including additional symptoms and their combinations.

The REal-time Assessment of Community Transmission-1 (REACT-1) study is a series of 2-3 weekly community-based cross-sectional surveys of SARS-CoV-2 PCR swab positivity carried out each month in England. It is based on random, non-overlapping samples of the general population aged 5 years and above.^8^ Here, we use REACT-1 data from rounds 2 to 7 (June to December 2020) and round 8 (January 2021) to evaluate the ability of (combinations of) reported symptoms in predicting PCR positivity in order to inform test allocation prioritisation. We compare the performances of models based on presence of at-least-one-of-four symptoms as per the current test allocation strategy, and parsimonious age-specific sets of symptoms found to be jointly predictive of PCR positivity. We also compare results across the most recent two rounds of data collection (round 7, November to December 2020, and round 8).

### Descriptive analyses of symptoms and PCR positivity

Of the 979,732 respondents completing a valid swab test who were enrolled in REACT-1 rounds 2–7, 19 were excluded due to missing information leaving 979,713 participants; of these, 870,875 (88.9%) reported no symptoms in the week prior to testing while 108,838 (11.1%) reported at least one of the 26 surveyed symptoms (Figure 1). We detected 4,169 PCR positives of whom 1,539 (36.9%) reported one or more symptoms in the past week (Figure 1-A and Supplementary Table 1).

**Figure 1.**
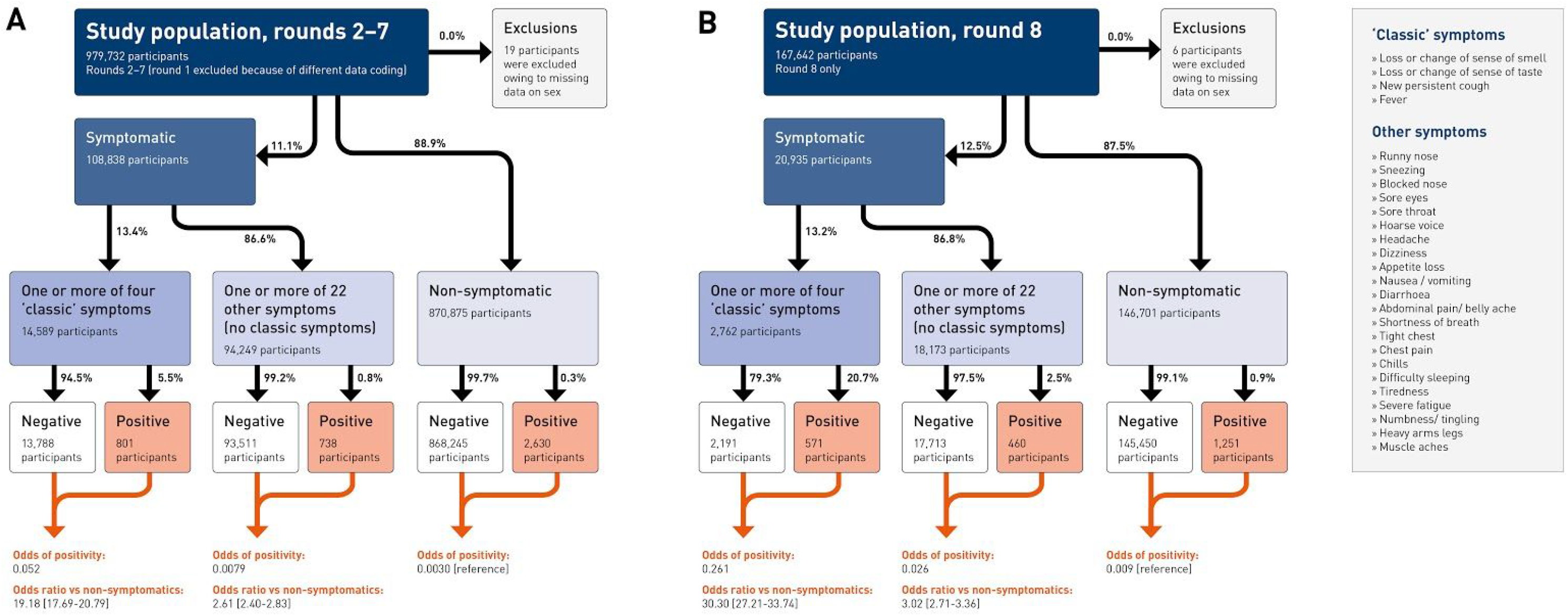
Flow chart showing numbers of participants, by symptom and PCR test status, included in A) rounds 2–7 and B) round 8 of the REACT-1 study.

In round 8 (Figure 1-B), after exclusion of 6 individuals with missing information, there were 167,636 participants, of whom 146,701 (87.5%) reported no symptoms in the past week and 20,935 (12.5%) reported at least one of the 26 surveyed symptoms. We detected 2,282 PCR positives of whom 1,031 (45.1%) reported one or more symptoms in the past week.

In rounds 2–7, we found a mean unweighted community prevalence of PCR positivity of 0.46%; 1.49% of participants reported at-least-one-of-four classic COVID-19 symptoms in the week before testing, increasing to 1.36% and 1.60% respectively in round 8. Odds ratio (OR) for testing positive among participants with at-least-one-of-four symptoms was 19.2 (95% confidence interval 17.7, 20.8) in rounds 2–7 and 30.3 (27.2, 33.7) in round 8 compared to people reporting no symptoms in the preceding week (Figure 1), consistent with results of the Office for National Statistics (ONS) Coronavirus (COVID-19) Infection Survey^9^. We calculated positive predictive values of 5.5% (rounds 2–7) and 20.7% (round 8) for at-least-one-of-four classic symptoms and 0.8% and 2.5%, respectively, for participants with one or more of the other 22 symptoms (Figure 1). In age-stratified analyses, we show that each of the 26 symptoms was associated with PCR positivity, except chest pain (rounds 2–7 and round 8) and shortness of breath (round 8 only) among 5–17 year olds (Supplementary Figure 1).

We constructed upset plots that show frequency of symptom occurrence or co-occurrence separately among PCR positives and negatives using the UpsetR package in R^10^ (Supplementary Figure 2). Among PCR positives, the most common symptom profiles were, in order: headache alone, co-occurring loss or change of sense of smell and taste, tiredness alone (rounds 2–7); and tiredness alone, headache alone, co-occurring loss or change of sense of smell and taste (round 8). Among PCR negatives, the most common symptom profiles were, in order: sore throat, headache, tiredness alone (rounds 2–7); and headache, tiredness, sore throat alone (round 8).

### Multivariate analyses

We then applied a stability selection procedure using penalised (Least Absolute Shrinkage and Selection Operator, LASSO) logistic regression among participants reporting at least one of the 26 surveyed symptoms. This was done separately at ages 5–17, 18–54 and 55+ years, to identify a model for each age group with parsimonious sets of symptoms jointly predictive of PCR positivity. We fitted and calibrated models on a 70% training set of data from rounds 2–7 and applied them in the remaining 30% as well as in all of round 8 data (see Methods). We considered stably selected symptoms to be predictive of PCR positivity, with selection proportions >50% across the N=1,000 models (based on 50% subsamples of the full study population). The selected symptoms and their penalised (shrunk) coefficients, averaged over models, are conditional on all other symptoms included.

In age-stratified analyses, six (5–17 years) or seven (18–54 and 55+ years) symptoms were jointly selected (Figure 2). All four classic symptoms (except new persistent cough for 5–17 years) were selected, along with chills in all age groups. Additional age-specific symptoms selected were headache (5–17 years), appetite loss (18–54 and 55+ years) and muscle aches (18–54 years) (positive coefficients) as well as runny nose (5–17 years) and numbness/tingling (55+ years) (negative coefficients). In contrast to results from univariate analyses (Supplementary Figure 1), runny nose appears to protect against risk of PCR positivity in the stability selection model for 5–17 year olds. This change in the sign of the association upon adjustment for other symptoms suggests that runny nose tends to co-occur with COVID-19 symptoms but is not specific to SARS-CoV-2 infection. Similarly, numbness/tingling reduces the predicted probability of getting a PCR positive test among 55+ year olds conditional on the other selected symptoms.

**Figure 2.**
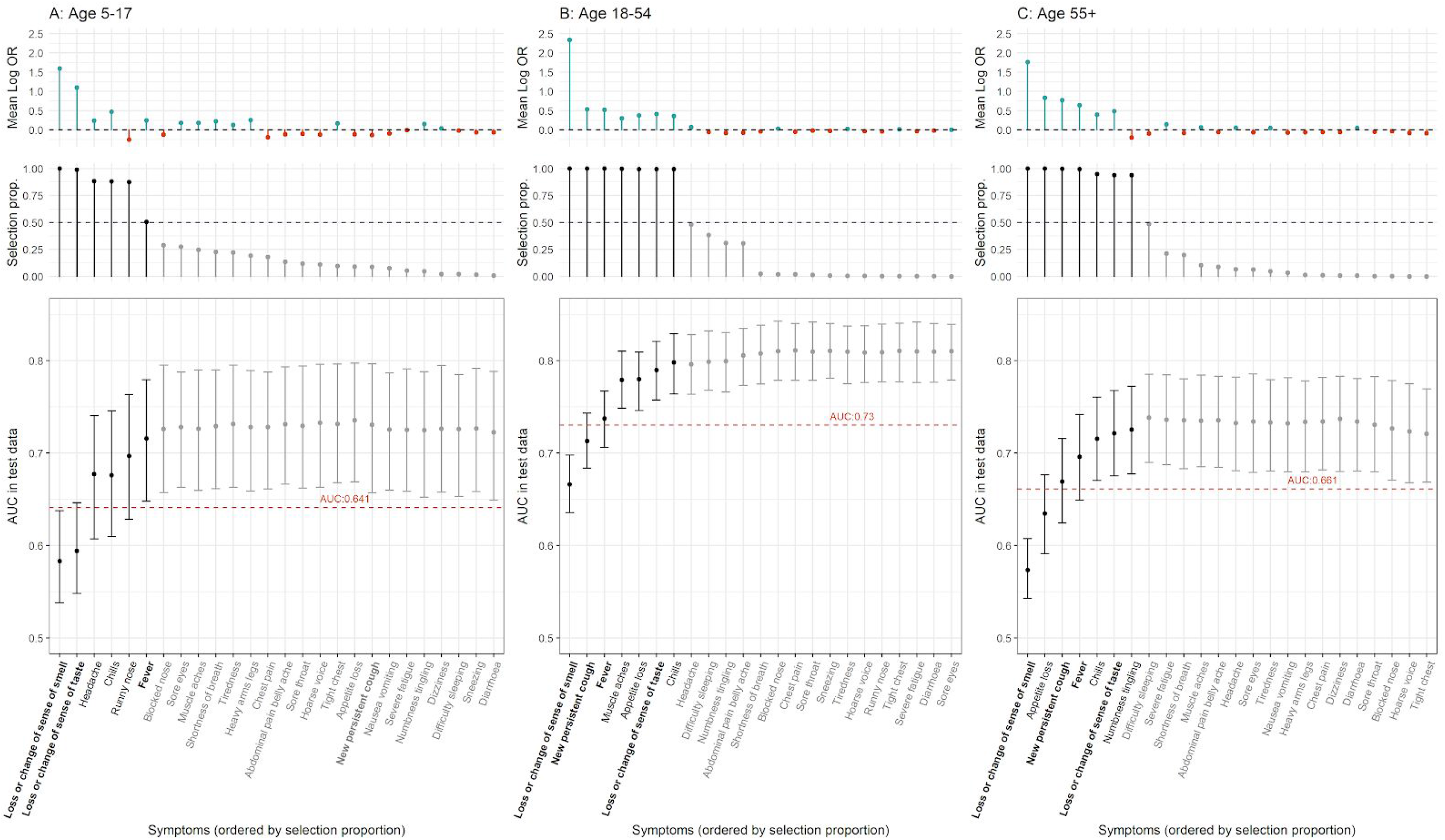
Results of LASSO stability selection using 1,000 models (with 50% subsamples of training data from rounds 2–7) at A) 5–17, B) 18–54 and C) 55+ years. Positive regression coefficients are presented in blue, and negative in red. Mean (penalised) regression coefficients across all models are shown in the top panel. Only symptoms selected at least once are displayed. The selection proportion for each symptom is shown in the middle panel, which is the proportion of 1,000 models that included the symptom; the horizontal dashed line represents the selection threshold of 50%. Symptoms are ordered according to their age-specific selection proportions, and the four classic COVID-19 symptoms are in bold. The bottom panel shows the area under the receiver operating characteristics curve (AUC) of models adding each variable to the model in order of selection proportion (from left to right). AUCs of the at-least-one-of-four symptoms testing strategy are displayed in red text (and red dashed line) on bottom panel plots.

In all age groups, addition of any other symptom (with non-zero selection proportion) made only incremental improvement to model performance (change in area under the receiver operating characteristic curve [AUC] < 0.02). Our stability selection models outperformed the at-least-one-of-four classic symptoms approach in rounds 2–7 test (hold-out) data with AUC = 0.72, 0.80, and 0.73 versus 0.64, 0.73 and 0.66 for 5–17, 18–54, and 55+ years, respectively (Figure 2). In sensitivity analyses, gradient-boosted trees,^11^ a non-linear alternative approach that accommodates complex interactions, did not outperform our penalised regression models in test data (Supplementary Figure 3).

### Round 7 versus round 8

We observed the highest prevalence of any REACT-1 round so far in round 8.^12^ We compared the prevalence of symptoms in round 7 and round 8 among PCR negatives and positives, separately, using a test-negative design.^13^ This was to account for possible seasonal effects that might explain the high rates of symptom reporting in round 8 (Supplementary Table 1 and Supplementary Table 2). Our analyses identified several symptoms with higher prevalence in round 8 than round 7, but, accounting for seasonal effects, prediction of PCR positivity only differed between rounds for loss or change of sense of smell (more predictive in round 7 than round 8) and (borderline) for new persistent cough, which was more predictive in round 8 (Supplementary Table 2). Our stability selection LASSO trained on round 7 and round 8 selected identical sets of variables jointly predictive of PCR positivity among individuals aged 55+ years, and slightly different sets of variables for the two younger age groups (Supplementary Figure 4). Despite these differences, the model trained on rounds 2–7 and tested on round 8 provided comparable predictive performance (AUC=0.78) to that tested on round 2–7 hold out data (AUC=0.77).

Our results may be relevant to understanding whether the symptom profile for variant B.1.1.7, which was first detected in England in September 2020, differs from previous strains. The rapid increase in the prevalence of this variant in England means that it became the dominant strain by January 2021, responsible for greater than 80% of infections at that time compared to ∼16% in mid-November 2020.^14^ Thus our findings of reduced PCR positivity risk associated with loss or change of sense of smell and increased risk for new persistent cough (in round 8 compared to round 7) may be due to variant B.1.1.7. These results are in keeping with findings from the ONS Coronavirus (COVID-19) Infection Survey where people testing PCR positive compatible with variant B.1.1.7 were less likely to report loss or change of taste or smell (both symptoms combined), and more likely to report cough, compared with earlier strains.^9^ Conversely, other differences in the symptoms associated with PCR positivity in round 7 and round 8 may, at least partially, be attributed to (possibly seasonal) changes in the prevalence of these symptoms.

### Community case detection

We show that at any proportion of community testing of symptomatic people, our models with selected symptoms would detect more cases than the at-least-one-of-four symptom approach currently adopted in England (Figure 3). If all people who reported at-least-one-of-four classic symptoms were tested (13% of the symptomatic population), we would detect 53% (rounds 2–7, Figure 3-A) and 55% (round 8, Figure 3-B) of symptomatic infections in the community; this would rise to 75% (76% in round 8) with optimised PCR testing allocation using our stability selection models (red dotted lines on Figure 3). Of these detected positives, 30% (rounds 2–7) and 27% (round 8) would not have reported any of the four classic symptoms, and so would not have been eligible for PCR testing. The ∼40% increase in case detection (above that for at-least-one-of-four symptoms) would involve a 2.7-fold increase in the number of tests allocated to the symptomatic population in England. Even so, only around one third of total community infections would be detected overall, due to the high proportion of non-symptomatic infections. This suggests that targeted testing of non-symptomatic individuals should also be considered alongside symptomatic testing^15^, for example, in areas of high prevalence.

**Figure 3.**
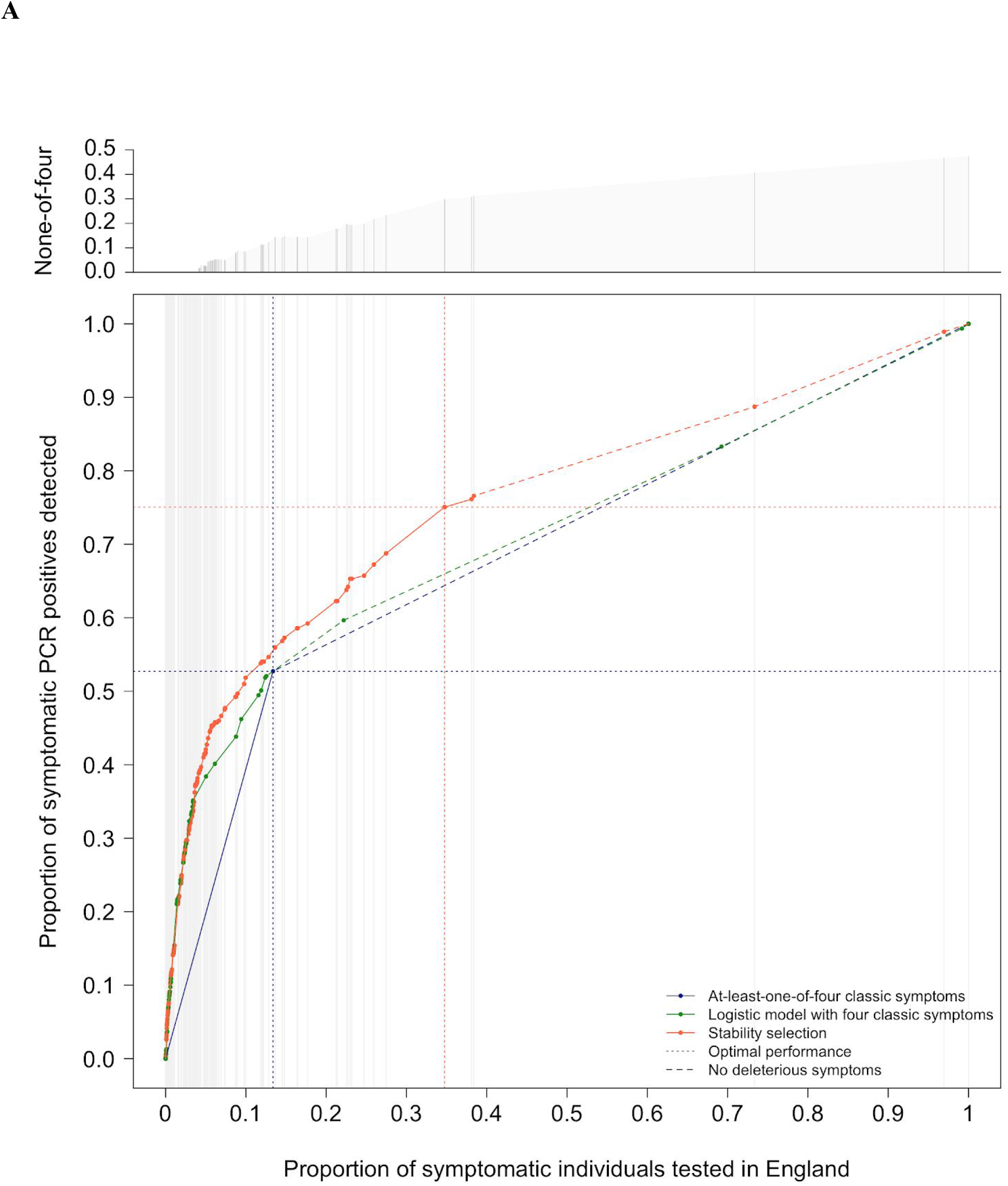
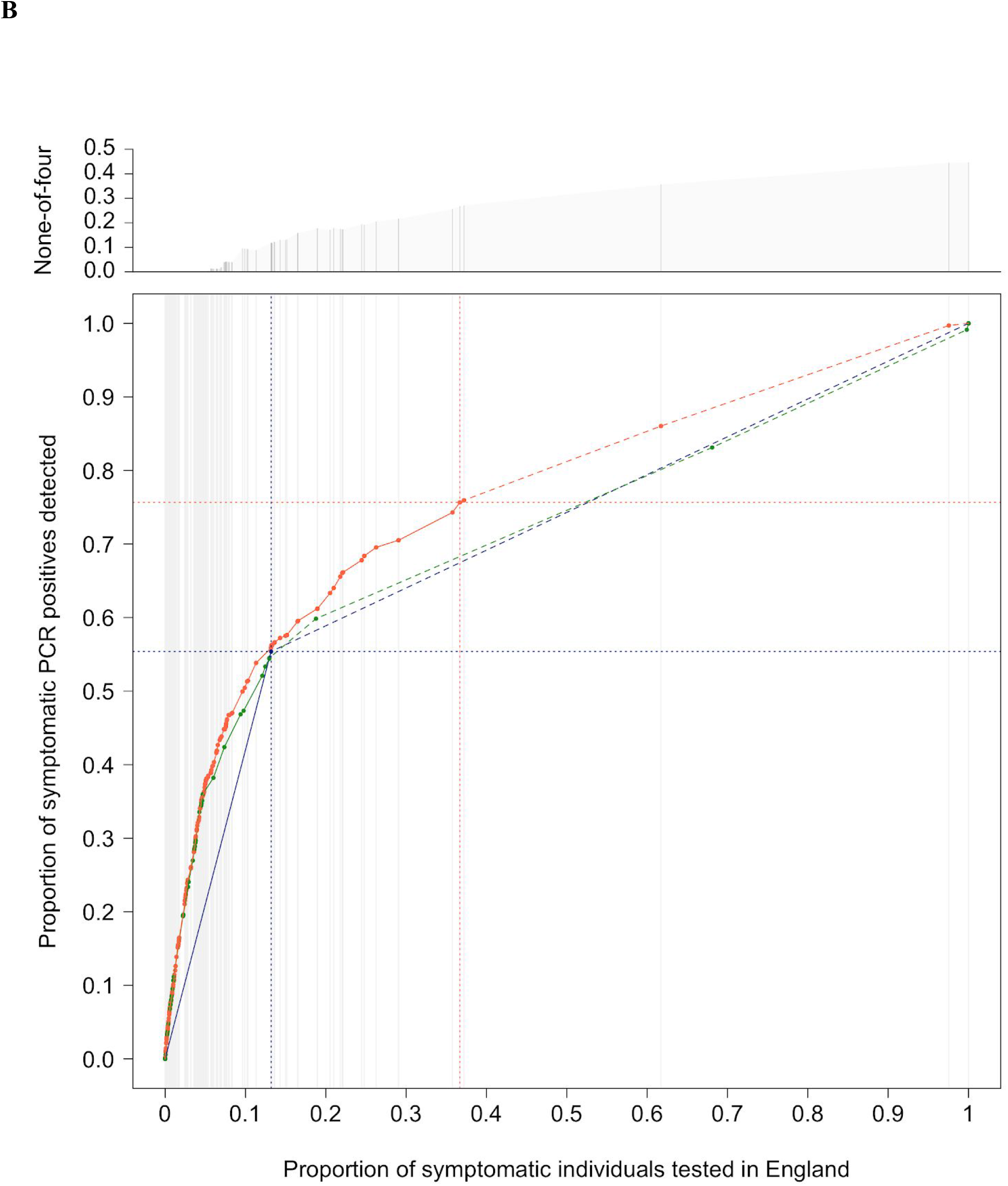
Simulation of test allocation and consequent proportions of symptomatic PCR positives (cases) detected, as a function of proportion of all symptomatic people tested, using models trained in rounds 2 to 7 and applied to A) hold-out test data (rounds 2 to 7) and B) round 8. Three scenarios are represented: (i) tests allocated to individuals with at-least-one-of-four classic symptoms (dark blue); tests allocated to the symptomatics with the highest probabilities of testing positive derived from (ii) the age-specific logistic regression models using the four classic symptoms as predictors (green); and (iii) the age-specific stability selection models (red). The dotted lines indicate the proportion of symptomatic individuals with at-least-one-of-four classic symptoms (dark blue), and an optimal proportion of symptomatic individuals eligible for a PCR testing based on prioritisation from the stability selection model (red). The points linked by dashed lines indicate individuals who would not have experienced (i) any of the four classic symptoms (dark blue, green), or (ii) any of the symptoms with positive coefficients identified by the stability selection models (red). The top panels shows the proportion of detected cases not reporting any of the four classic symptoms.

## Discussion

Previous models of the relationship between symptoms and infection identified fatigue and skipping meals in addition to the classic four symptoms as predictive of reported PCR positivity, although these symptoms did not appear to vary by age.^16^ In contrast, we show here that symptom profile may differ by age, and that age-stratified symptom-based models may offer improved predictions of PCR positivity. For every possible number of PCR tests done, more COVID-19 cases would be identified by our models than with the current at-least-one-of-four classic symptoms strategy.

We note that the selected symptoms all contribute to prediction of PCR positivity: thus, for example, loss or change of sense of taste and loss or change of sense of smell confer independent predictive information and therefore both symptoms should be considered. Likewise, reporting chills provides additional information at all ages beyond fever, and loss of appetite (among adults) adds additional information over and above loss or change of sense of taste or smell. Beyond the symptoms selected by LASSO stability selection, however, we found no meaningful improvement in prediction of PCR positivity, indicating that our selection procedure provided an informative but parsimonious set of variables.

Limitations of our study include that the participants, though randomly selected from the community, may not be fully representative and therefore results may not be directly applicable to the whole population. However, our sampling procedure, which provided approximately equal numbers of participants in all 315 lower tier local authority areas in England, ensured wide geographical coverage and captured the socio-demographic and ethnic diversity of the population of England. In addition, self-administered questionnaires are subject to possible recall and other biases and therefore may not give an accurate representation of the symptom profiles among the population as a whole. Also, as sampling in each round was cross-sectional, some individuals may have been infected (and had symptoms) more than one week before the swab was obtained, but were no longer symptomatic at the time of the study.

Our models with stably selected symptoms have the advantage of increasing the yield of positive tests leading to greater numbers of infected people being required to self-isolate, and therefore reducing the pool of infection in the community. However, this may increase the need to support individuals who may be economically disadvantaged by having to self-isolate. Also, since a high proportion of community infections are non-symptomatic, the majority of infected individuals would still go undetected. Thus, at the same time as widening the availability of testing among symptomatic people, consideration should be given to targeted testing of non-symptomatic individuals, for example, those in areas of high prevalence.

In summary, we show that the yield for community COVID-19 case detection could be increased. This would involve inclusion of a few additional symptoms in the criteria for test eligibility, in particular, chills, headache, appetite loss and muscle aches.

## Methods

### Study Population

REACT-1 is a series of community prevalence surveys of SARS-CoV-2 virus swab-positivity in England, conducted at approximately monthly intervals since May 2020. In each round of the study, recruitment letters are sent at random to a nationally representative sample of individuals aged 5 years and over using the National Health Service patient register across the 315 lower-tier local authorities in England. Each round aims to gather around 160,000 valid responses. Data from round 1 were excluded from our analysis as the symptom questions asked in that round were not consistent with subsequent rounds.^8^

### Statistical analyses

#### Univariate models

Within each age group we used logistic regressions to model the risk of testing positive for SARS-CoV-2 infection, conditionally on having experienced any of 26 symptoms in the week prior to testing. We report ORs from such univariate models for each of the 26 symptoms separately.

### Multivariate analyses

We adopted a variable selection approach to account for correlation and co-occurrence patterns across the 26 symptoms, and to model their joint and mutually adjusted effects on risk of PCR positivity. We used stability selection applied to LASSO (Least Absolute Shrinkage and Selection Operator) penalised logistic regression including all 26 symptoms as predictors and PCR test positive/negative as the outcome variable.^17^ In our stability selection, LASSO models are refitted on 1,000 random subsamples of the data, with a representative proportion of negative and positive tests retained in each subsample. As a measure of importance of each symptom in predicting PCR positivity, conditional on the other symptoms, we report the number of times each symptom was included in the model across the 1,000 subsamples. We considered symptoms with selection proportion above 50% as stable and included them in the stability selection model. The penalty parameter is calibrated to ensure that fewer than five predictors could be falsely selected in this model (per family error rate). For stable symptoms, the average ORs (conditionally on selection in the refitted LASSO models with calibrated penalty) are computed and used as effect size estimates in the stability selection model.

As a non-parametric alternative, we used gradient-boosted forests, as implemented in XGBoost.^11^ Model complexity was calibrated by tuning the number of trees (‘nrounds’) and tree depth (‘max_depth’) hyperparameters using 5-fold cross-validation, within each age group. Other hyperparameters were set to default.

For both multivariate models, the rounds 2–7 dataset (restricted to people reporting any of the 26 symptoms) was split into a 70% training set (76,187 observations, of which 1,078 were positive) and a 30% test set (32,651 observations, of which 461 were positive). All models were trained on the 70% training set and evaluated on the test set, as well as on round 8 data.

#### Evaluation of the test allocation strategy

For each participant in the test set, we used age-specific effect size estimates to derive an individual predicted probability of testing positive from (i) the stability selection model, (ii) the (unpenalised) logistic model with the four classic symptoms, and (iii) the currently implemented strategy (probability of 1 if experiencing any of the four main symptoms, and zero otherwise). To compare the performances of these three symptom-based strategies to select individuals for PCR testing, we computed the AUC for each.

We also simulated reallocating PCR tests in the community on the basis of model-derived risk scores and with the current testing strategy (at-least-one-of-four classic symptoms). We reported the proportion of symptomatic COVID-19 cases that would be detected (on PCR testing) using each of the three strategies for prioritisation, as a function of the proportion of symptomatic participants tested.

#### Round 7 versus round 8

We adopted a similar stability selection logistic LASSO approach on data restricted to participants from rounds 7 and 8 separately, and compared resulting selection proportions. To account for seasonal effects on the prevalence of symptoms, irrespective of PCR positivity, we adopted a test-negative design approach comparing separately among PCR positives and negatives across rounds. We investigated differences in the ORs (measuring the association between the symptom and PCR positivity) between round 7 and round 8 using a logistic model setting the symptom as outcome, the round and PCR positivity as predictors, and including a multiplicative interaction term. The effect of round is independent of PCR positivity (and can be interpreted as the effect of season on the symptom’s prevalence), the effect of PCR positivity measures the association between symptoms prevalence and the outcome of the test, irrespective of the round, and the interaction term measures a possible change in the association between PCR positivity and the symptoms, independently of the effect of season.

## Data Availability

Summary statistics and descriptive tables are available in Supplementary information; R scripts used to perform the analyses can be downloaded at https://github.com/mathzero/react_symptom_risk_prediciton.

## Ethical Approval

We obtained research ethics approval from the South Central-Berkshire B Research Ethics Committee (IRAS ID: 283787).

## Contributors

JE, MW, and BB are joint first authors. MC-H and PE are joint last authors. JE, MW, MC-H, BB and PE conceived the study and drafted the manuscript. JE, BB, and MDW performed the statistical analyses. SR, HW and GC provided insights into the study design and results interpretation. AD and PE obtained funding. All authors revised the manuscript for important intellectual content and approved the submission of the manuscript. MC-H and PE had full access to the data and take responsibility for the integrity of the data and the accuracy of the data analysis and for the decision to submit for publication.

## Acknowledgements

We thank key collaborators on this work – Ipsos MORI: Kelly Beaver, Sam Clemens, Gary Welch, Nicholas Gilby, Kelly Ward and Kevin Pickering; Institute of Global Health Innovation at Imperial College: Gianluca Fontana, Dr Hutan Ashrafian, Sutha Satkunarajah, Didi Thompson, Justine Alford and Lenny Naar; Molecular Diagnostic Unit, Imperial College London: Prof. Graham Taylor; North West London Pathology and Public Health England for help in calibration of the laboratory analyses; NHS Digital for access to the NHS register; and the Department of Health and Social Care for logistic support.

## Data sharing statement

Summary statistics and descriptive tables are available in Supplementary information; R scripts used to perform the analyses can be downloaded at https://github.com/barbarabodinier/Community_detection_COVID-19.

## Supplementary information

**Supplementary Table 1.** Symptom prevalence by age group among SARS-CoV-2 PCR test negative and positive respondents in REACT-1 A) rounds 2 to 7 and B) round 8. **A** Rounds 2 to 7

|  | Age 5-17 |  | Age 18-54 |  | Age 55+ |  |
| --- | --- | --- | --- | --- | --- | --- |
| *Symptom* | *Negative* | *Positive* | *Negative* | *Positive* | *Negative* | *Positive* |
| Full cohort | 127838 | 704 | 417631 | 2064 | 430075 | 1401 |
| Any one of four Classic symptoms | 2130 (1.7%) | 87 (12.4%) | 7446 (1.8%) | 497 (24.1%) | 4212 (1%) | 217 (15.5%) |
| Loss of sense of smell | 253 (0.2%) | 40 (5.7%) | 1365 (0.3%) | 309 (15%) | 689 (0.2%) | 90 (6.4%) |
| Loss of sense of taste | 244 (0.2%) | 33 (4.7%) | 1584 (0.4%) | 265 (12.8%) | 937 (0.2%) | 90 (6.4%) |
| New persistent cough | 1148 (0.9%) | 23 (3.3%) | 2958 (0.7%) | 184 (8.9%) | 1492 (0.3%) | 76 (5.4%) |
| Fever | 985 (0.8%) | 41 (5.8%) | 3403 (0.8%) | 195 (9.4%) | 1971 (0.5%) | 98 (7%) |
| Runny nose | 5823 (4.6%) | 68 (9.7%) | 15719 (3.8%) | 266 (12.9%) | 8154 (1.9%) | 123 (8.8%) |
| Sneezing | 4547 (3.6%) | 67 (9.5%) | 15595 (3.7%) | 281 (13.6%) | 8490 (2%) | 130 (9.3%) |
| Blocked nose | 5126 (4%) | 71 (10.1%) | 11954 (2.9%) | 299 (14.5%) | 5465 (1.3%) | 88 (6.3%) |
| Sore eyes | 1083 (0.8%) | 28 (4%) | 7800 (1.9%) | 161 (7.8%) | 6301 (1.5%) | 79 (5.6%) |
| Sore throat | 4792 (3.7%) | 76 (10.8%) | 18702 (4.5%) | 306 (14.8%) | 8184 (1.9%) | 103 (7.4%) |
| Hoarse voice | 1384 (1.1%) | 17 (2.4%) | 4621 (1.1%) | 133 (6.4%) | 3507 (0.8%) | 61 (4.4%) |
| Headache | 4227 (3.3%) | 106 (15.1%) | 26226 (6.3%) | 540 (26.2%) | 12366 (2.9%) | 200 (14.3%) |
| Dizziness | 1042 (0.8%) | 25 (3.6%) | 7502 (1.8%) | 168 (8.1%) | 5118 (1.2%) | 69 (4.9%) |
| Appetite loss | 1193 (0.9%) | 25 (3.6%) | 3887 (0.9%) | 194 (9.4%) | 2481 (0.6%) | 115 (8.2%) |
| Nausea/vomiting | 1267 (1%) | 25 (3.6%) | 5466 (1.3%) | 112 (5.4%) | 2566 (0.6%) | 42 (3%) |
| Diarrhoea | 883 (0.7%) | 16 (2.3%) | 6949 (1.7%) | 116 (5.6%) | 3978 (0.9%) | 65 (4.6%) |
| Abdominal pain / belly ache | 2386 (1.9%) | 33 (4.7%) | 8986 (2.2%) | 120 (5.8%) | 5541 (1.3%) | 61 (4.4%) |
| Shortness of breath | 575 (0.4%) | 18 (2.6%) | 6392 (1.5%) | 146 (7.1%) | 6292 (1.5%) | 77 (5.5%) |
| Tight chest | 508 (0.4%) | 16 (2.3%) | 5344 (1.3%) | 146 (7.1%) | 3429 (0.8%) | 58 (4.1%) |
| Chest pain | 387 (0.3%) | 5 (0.7%) | 2582 (0.6%) | 61 (3%) | 1493 (0.3%) | 22 (1.6%) |
| Chills | 904 (0.7%) | 40 (5.7%) | 4199 (1%) | 212 (10.3%) | 2569 (0.6%) | 97 (6.9%) |
| Difficulty sleeping | 1738 (1.4%) | 33 (4.7%) | 15654 (3.7%) | 242 (11.7%) | 10548 (2.5%) | 108 (7.7%) |
| Tiredness | 2884 (2.3%) | 70 (9.9%) | 24240 (5.8%) | 482 (23.4%) | 14930 (3.5%) | 246 (17.6%) |
| Severe fatigue | 271 (0.2%) | 7 (1%) | 2831 (0.7%) | 99 (4.8%) | 1697 (0.4%) | 51 (3.6%) |
| Numbness/tingling | 276 (0.2%) | 7 (1%) | 4917 (1.2%) | 77 (3.7%) | 4721 (1.1%) | 33 (2.4%) |
| Heavy arms/legs | 353 (0.3%) | 13 (1.8%) | 4556 (1.1%) | 136 (6.6%) | 3775 (0.9%) | 61 (4.4%) |
| Muscle aches | 1163 (0.9%) | 34 (4.8%) | 11741 (2.8%) | 378 (18.3%) | 9052 (2.1%) | 172 (12.3%) |

B Round 8
|  | Age 5-17 |  | Age 18-54 |  | Age 55+ |  |
| --- | --- | --- | --- | --- | --- | --- |
| *Symptom* | *Negative* | *Positive* | *Negative* | *Positive* | *Negative* | *Positive* |
| Full cohort | 20331 | 342 | 70380 | 1145 | 74643 | 795 |
| Any one of four Classic symptoms | 146 (0.7%) | 36 (10.5%) | 1280 (1.8%) | 359 (31.4%) | 765 (1%) | 176 (22.1%) |
| Loss of sense of smell | 30 (0.1%) | 19 (5.6%) | 296 (0.4%) | 193 (16.9%) | 161 (0.2%) | 68 (8.6%) |
| Loss of sense of taste | 21 (0.1%) | 16 (4.7%) | 317 (0.5%) | 176 (15.4%) | 193 (0.3%) | 83 (10.4%) |
| New persistent cough | 39 (0.2%) | 13 (3.8%) | 545 (0.8%) | 167 (14.6%) | 301 (0.4%) | 81 (10.2%) |
| Fever | 76 (0.4%) | 9 (2.6%) | 478 (0.7%) | 142 (12.4%) | 312 (0.4%) | 62 (7.8%) |
| Runny nose | 320 (1.6%) | 33 (9.6%) | 2997 (4.3%) | 189 (16.5%) | 1917 (2.6%) | 101 (12.7%) |
| Sneezing | 292 (1.4%) | 26 (7.6%) | 2672 (3.8%) | 199 (17.4%) | 1938 (2.6%) | 99 (12.5%) |
| Blocked nose | 330 (1.6%) | 34 (9.9%) | 2160 (3.1%) | 212 (18.5%) | 1230 (1.6%) | 84 (10.6%) |
| Sore eyes | 133 (0.7%) | 19 (5.6%) | 1404 (2%) | 108 (9.4%) | 1075 (1.4%) | 50 (6.3%) |
| Sore throat | 247 (1.2%) | 31 (9.1%) | 2885 (4.1%) | 212 (18.5%) | 1485 (2%) | 77 (9.7%) |
| Hoarse voice | 43 (0.2%) | 7 (2%) | 695 (1%) | 80 (7%) | 567 (0.8%) | 59 (7.4%) |
| Headache | 557 (2.7%) | 50 (14.6%) | 5234 (7.4%) | 345 (30.1%) | 2519 (3.4%) | 158 (19.9%) |
| Dizziness | 150 (0.7%) | 11 (3.2%) | 1307 (1.9%) | 107 (9.3%) | 894 (1.2%) | 64 (8.1%) |
| Appetite loss | 173 (0.9%) | 15 (4.4%) | 713 (1%) | 136 (11.9%) | 473 (0.6%) | 112 (14.1%) |
| Nausea/vomiting | 144 (0.7%) | 7 (2%) | 928 (1.3%) | 70 (6.1%) | 448 (0.6%) | 32 (4%) |
| Diarrhoea | 96 (0.5%) | 7 (2%) | 1138 (1.6%) | 85 (7.4%) | 666 (0.9%) | 46 (5.8%) |
| Abdominal pain / belly ache | 292 (1.4%) | 12 (3.5%) | 1550 (2.2%) | 74 (6.5%) | 1009 (1.4%) | 62 (7.8%) |
| Shortness of breath | 76 (0.4%) | 3 (0.9%) | 1287 (1.8%) | 119 (10.4%) | 1159 (1.6%) | 50 (6.3%) |
| Tight chest | 68 (0.3%) | 4 (1.2%) | 1137 (1.6%) | 115 (10%) | 726 (1%) | 45 (5.7%) |
| Chest pain | 37 (0.2%) | 2 (0.6%) | 533 (0.8%) | 54 (4.7%) | 267 (0.4%) | 18 (2.3%) |
| Chills | 109 (0.5%) | 11 (3.2%) | 1220 (1.7%) | 165 (14.4%) | 836 (1.1%) | 92 (11.6%) |
| Difficulty sleeping | 301 (1.5%) | 20 (5.8%) | 3341 (4.7%) | 162 (14.1%) | 2193 (2.9%) | 88 (11.1%) |
| Tiredness | 347 (1.7%) | 37 (10.8%) | 4339 (6.2%) | 339 (29.6%) | 2557 (3.4%) | 183 (23%) |
| Severe fatigue | 42 (0.2%) | 6 (1.8%) | 567 (0.8%) | 82 (7.2%) | 313 (0.4%) | 46 (5.8%) |
| Numbness/tingling | 28 (0.1%) | 5 (1.5%) | 865 (1.2%) | 44 (3.8%) | 793 (1.1%) | 25 (3.1%) |
| Heavy arms/legs | 41 (0.2%) | 6 (1.8%) | 792 (1.1%) | 98 (8.6%) | 627 (0.8%) | 52 (6.5%) |
| Muscle aches | 112 (0.6%) | 19 (5.6%) | 2163 (3.1%) | 228 (19.9%) | 1536 (2.1%) | 119 (15%) |

**Supplementary Table 2.**
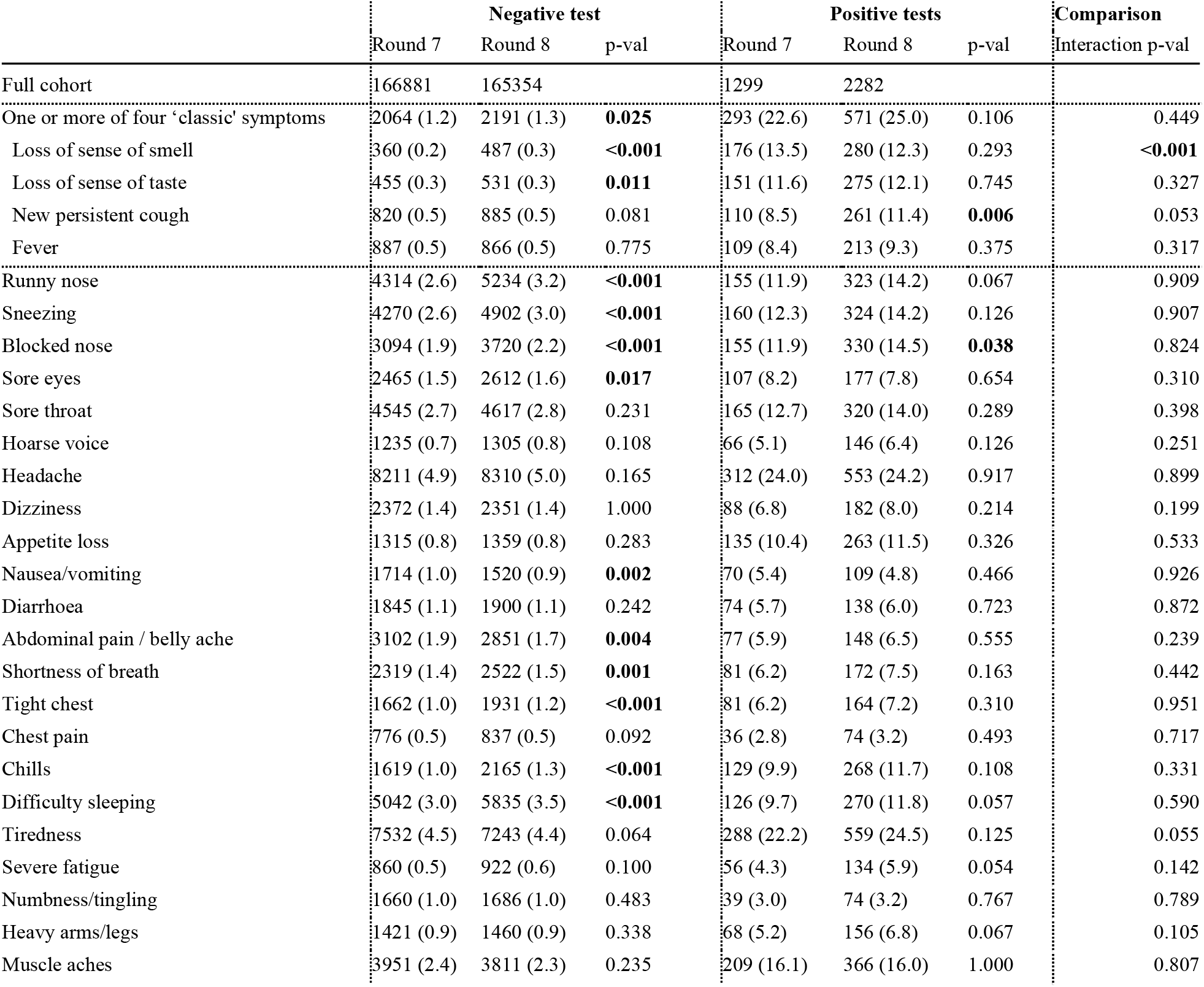
Comparison of prevalence of symptoms in test negatives and test positives in round 7 vs round 8. P-values from Chi^2^ test measure the differences in prevalence across rounds in test negatives and test positives separately. The comparison was performed using a logistic model regressing the symptom status against round, test result and a multiplicative interaction term between these two predictors. The interaction term measures a possible change in the association between PCR positivity and the symptoms, independently of the effect of season, and we report the corresponding p-value.

|  | Negative test |  |  | Positive tests |  |  | Comparison |
| --- | --- | --- | --- | --- | --- | --- | --- |
|  | Round 7 | Round 8 | p-val | Round 7 | Round 8 | p-val | Interaction p-val |
| Full cohort | 166881 | 165354 |  | 1299 | 2282 |  |  |
| One or more of four 'classic' symptoms | 2064 (1.2) | 2191 (1.3) | **0.025** | 293 (22.6) | 571 (25.0) | 0.106 | 0.449 |
| Loss of sense of smell | 360 (0.2) | 487 (0.3) | **<0.001** | 176 (13.5) | 280 (12.3) | 0.293 | **<0.001** |
| Loss of sense of taste | 455 (0.3) | 531 (0.3) | **0.011** | 151 (11.6) | 275 (12.1) | 0.745 | 0.327 |
| New persistent cough | 820 (0.5) | 885 (0.5) | 0.081 | 110 (8.5) | 261 (11.4) | **0.006** | 0.053 |
| Fever | 887 (0.5) | 866 (0.5) | 0.775 | 109 (8.4) | 213 (9.3) | 0.375 | 0.317 |
| Runny nose | 4314 (2.6) | 5234 (3.2) | **<0.001** | 155 (11.9) | 323 (14.2) | 0.067 | 0.909 |
| Sneezing | 4270 (2.6) | 4902 (3.0) | **<0.001** | 160 (12.3) | 324 (14.2) | 0.126 | 0.907 |
| Blocked nose | 3094 (1.9) | 3720 (2.2) | **<0.001** | 155 (11.9) | 330 (14.5) | **0.038** | 0.824 |
| Sore eyes | 2465 (1.5) | 2612 (1.6) | **0.017** | 107 (8.2) | 177 (7.8) | 0.654 | 0.310 |
| Sore throat | 4545 (2.7) | 4617 (2.8) | 0.231 | 165 (12.7) | 320 (14.0) | 0.289 | 0.398 |
| Hoarse voice | 1235 (0.7) | 1305 (0.8) | 0.108 | 66 (5.1) | 146 (6.4) | 0.126 | 0.251 |
| Headache | 8211 (4.9) | 8310 (5.0) | 0.165 | 312 (24.0) | 553 (24.2) | 0.917 | 0.899 |
| Dizziness | 2372 (1.4) | 2351 (1.4) | 1.000 | 88 (6.8) | 182 (8.0) | 0.214 | 0.199 |
| Appetite loss | 1315 (0.8) | 1359 (0.8) | 0.283 | 135 (10.4) | 263 (11.5) | 0.326 | 0.533 |
| Nausea/vomiting | 1714 (1.0) | 1520 (0.9) | **0.002** | 70 (5.4) | 109 (4.8) | 0.466 | 0.926 |
| Diarrhoea | 1845 (1.1) | 1900 (1.1) | 0.242 | 74 (5.7) | 138 (6.0) | 0.723 | 0.872 |
| Abdominal pain / belly ache | 3102 (1.9) | 2851 (1.7) | **0.004** | 77 (5.9) | 148 (6.5) | 0.555 | 0.239 |
| Shortness of breath | 2319 (1.4) | 2522 (1.5) | **0.001** | 81 (6.2) | 172 (7.5) | 0.163 | 0.442 |
| Tight chest | 1662 (1.0) | 1931 (1.2) | **<0.001** | 81 (6.2) | 164 (7.2) | 0.310 | 0.951 |
| Chest pain | 776 (0.5) | 837 (0.5) | 0.092 | 36 (2.8) | 74 (3.2) | 0.493 | 0.717 |
| Chills | 1619 (1.0) | 2165 (1.3) | **<0.001** | 129 (9.9) | 268 (11.7) | 0.108 | 0.331 |
| Difficulty sleeping | 5042 (3.0) | 5835 (3.5) | **<0.001** | 126 (9.7) | 270 (11.8) | 0.057 | 0.590 |
| Tiredness | 7532 (4.5) | 7243 (4.4) | 0.064 | 288 (22.2) | 559 (24.5) | 0.125 | 0.055 |
| Severe fatigue | 860 (0.5) | 922 (0.6) | 0.100 | 56 (4.3) | 134 (5.9) | 0.054 | 0.142 |
| Numbness/tingling | 1660 (1.0) | 1686 (1.0) | 0.483 | 39 (3.0) | 74 (3.2) | 0.767 | 0.789 |
| Heavy arms/legs | 1421 (0.9) | 1460 (0.9) | 0.338 | 68 (5.2) | 156 (6.8) | 0.067 | 0.105 |
| Muscle aches | 3951 (2.4) | 3811 (2.3) | 0.235 | 209 (16.1) | 366 (16.0) | 1.000 | 0.807 |

**Supplementary Figure 1.**
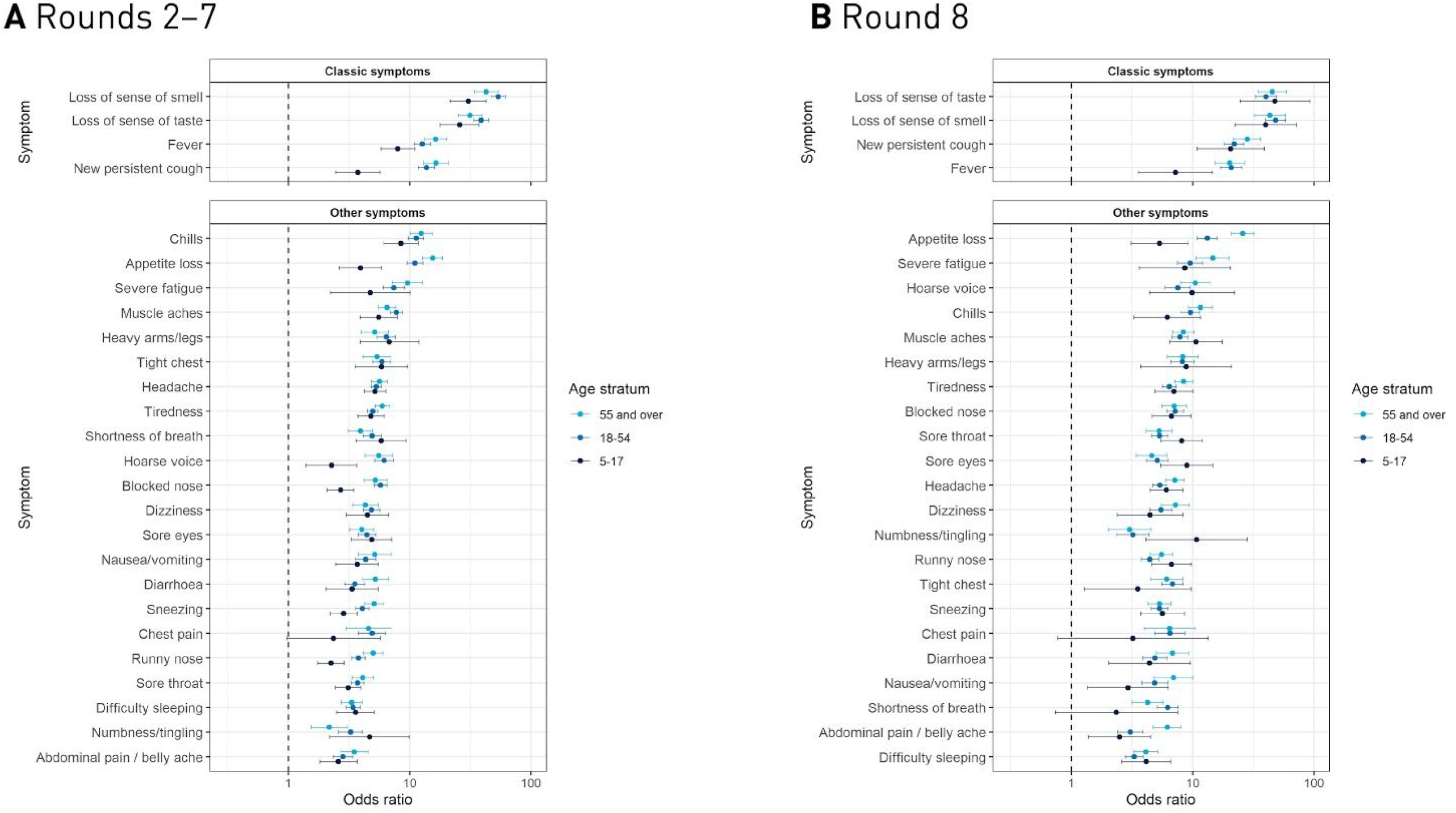
Results from univariate logistic regression models of PCR positivity for each of the 26 symptoms, separately. Effect size estimates are expressed as odds ratios (with 95% confidence intervals) and presented for 5–17 years (dark blue), 18–54 years (blue), and 55+ years (cyan) age groups in A) rounds 2–7 and B) round 8.

**Supplementary Figure 2.**
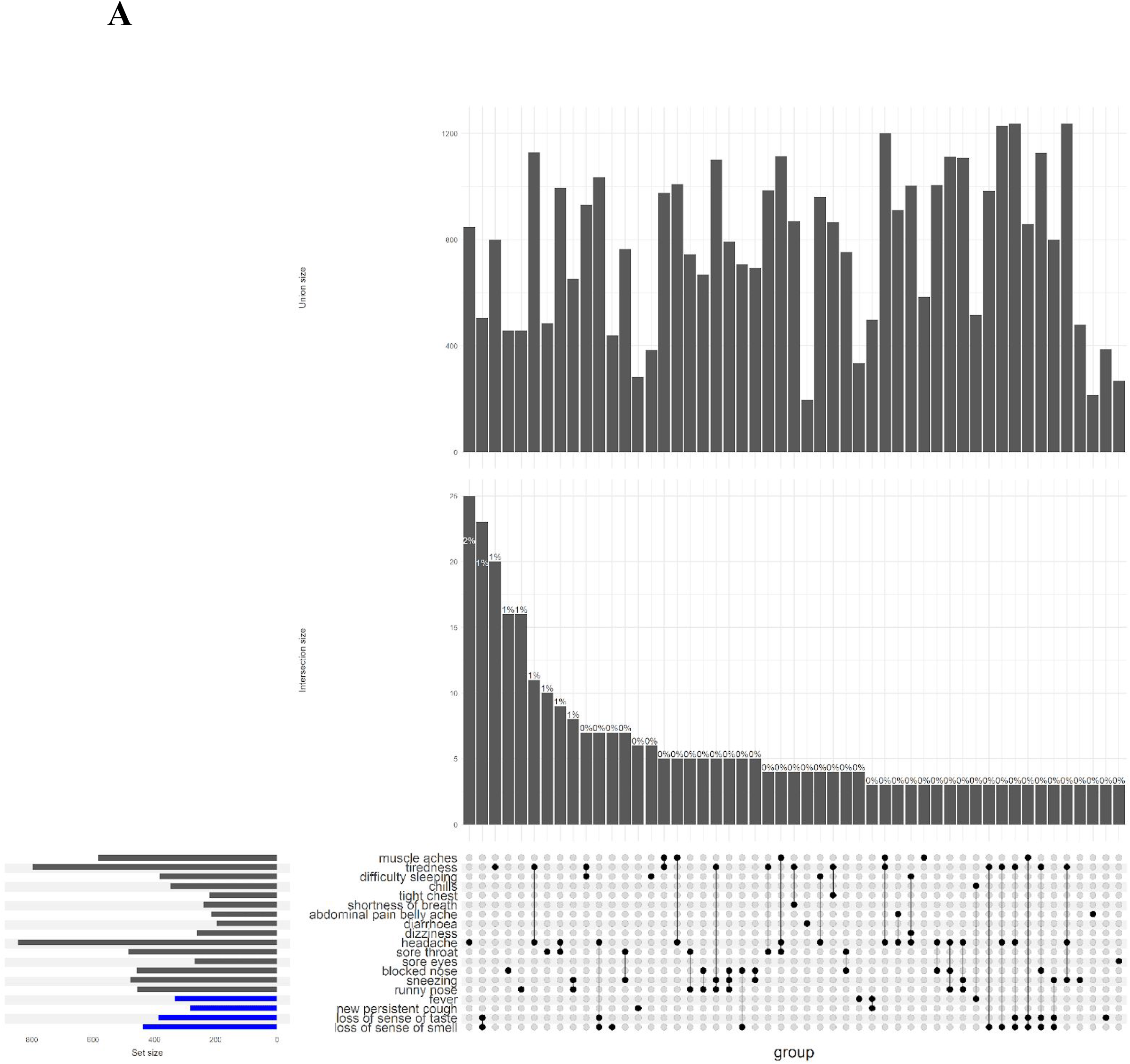
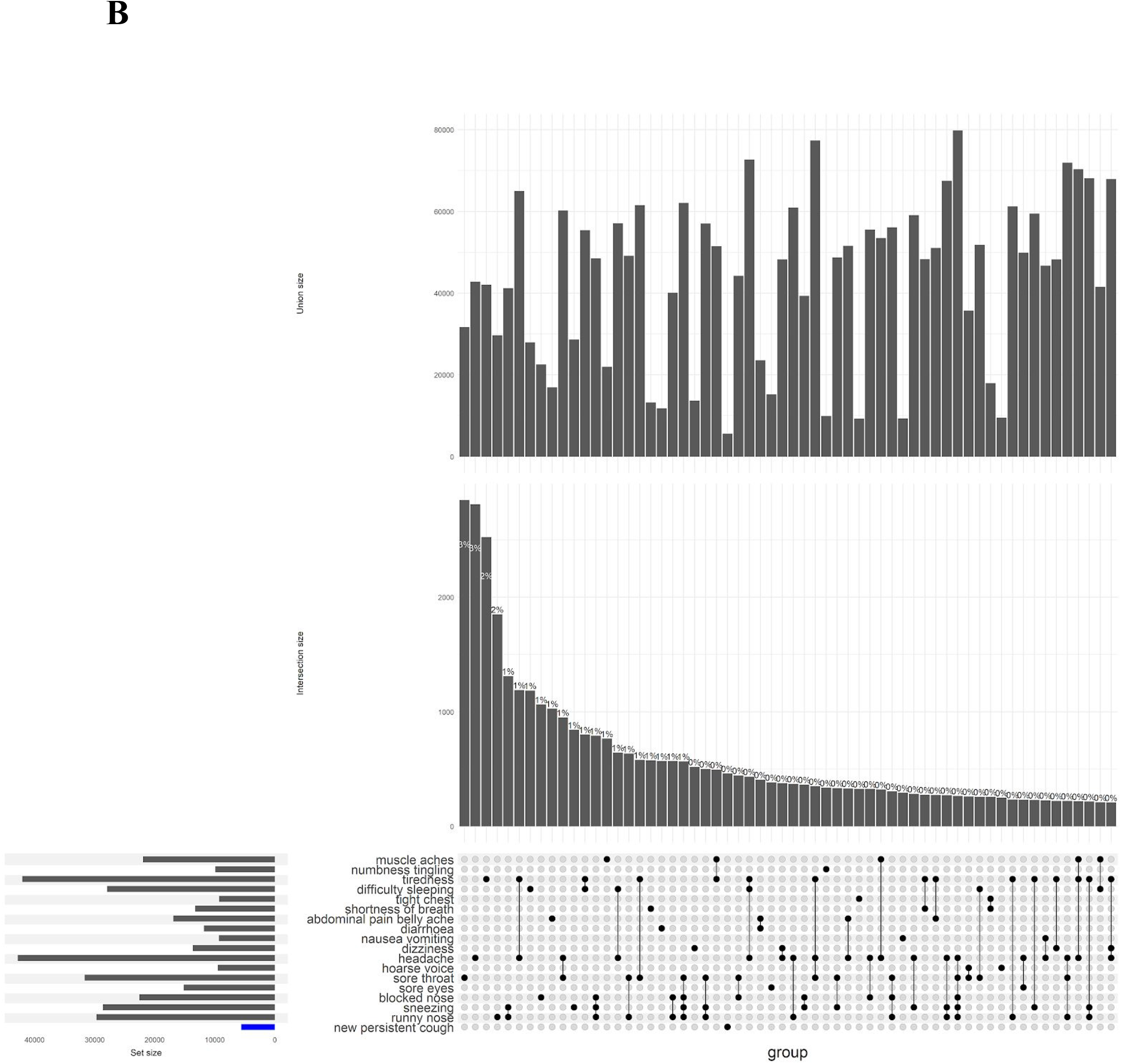
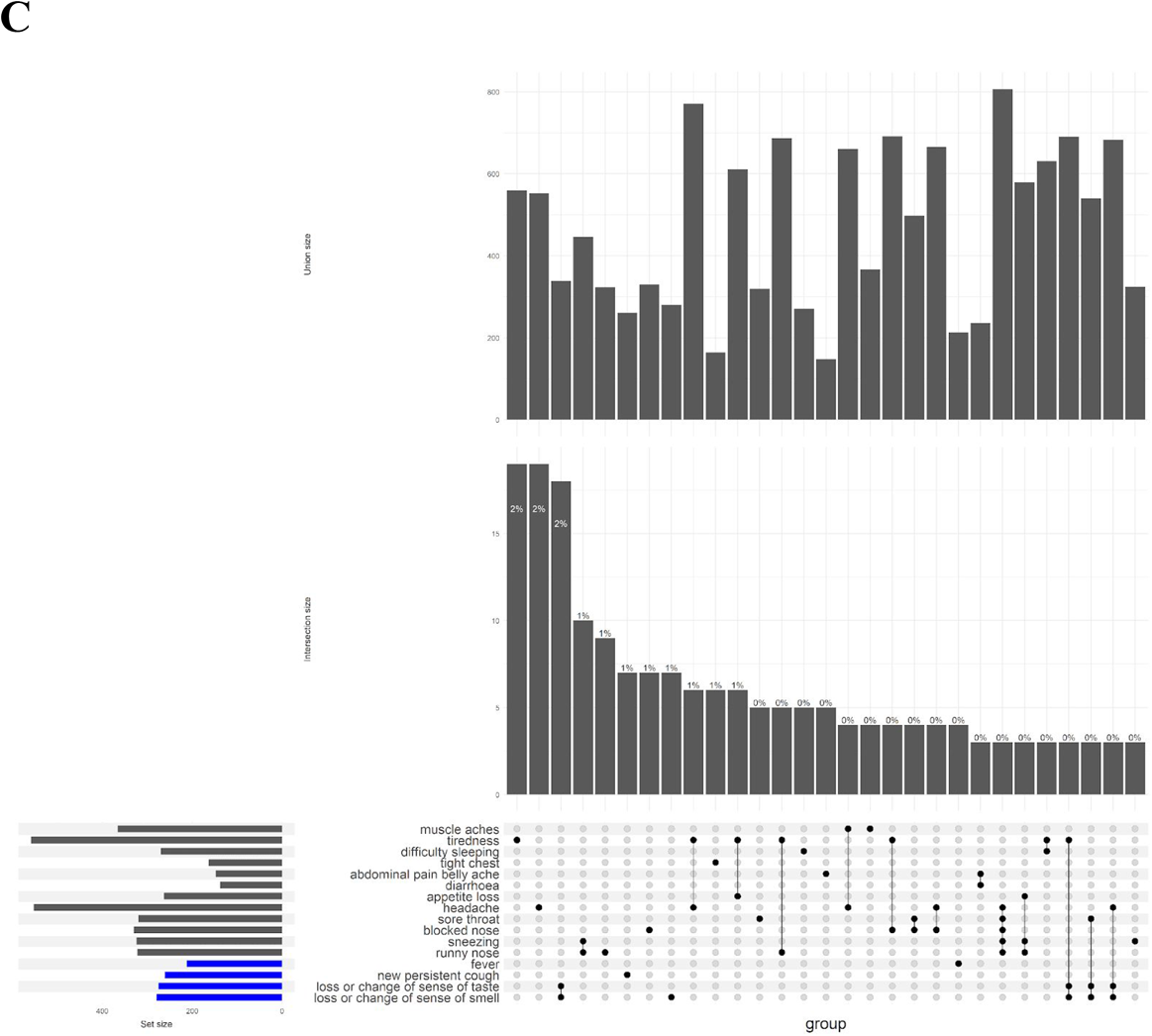
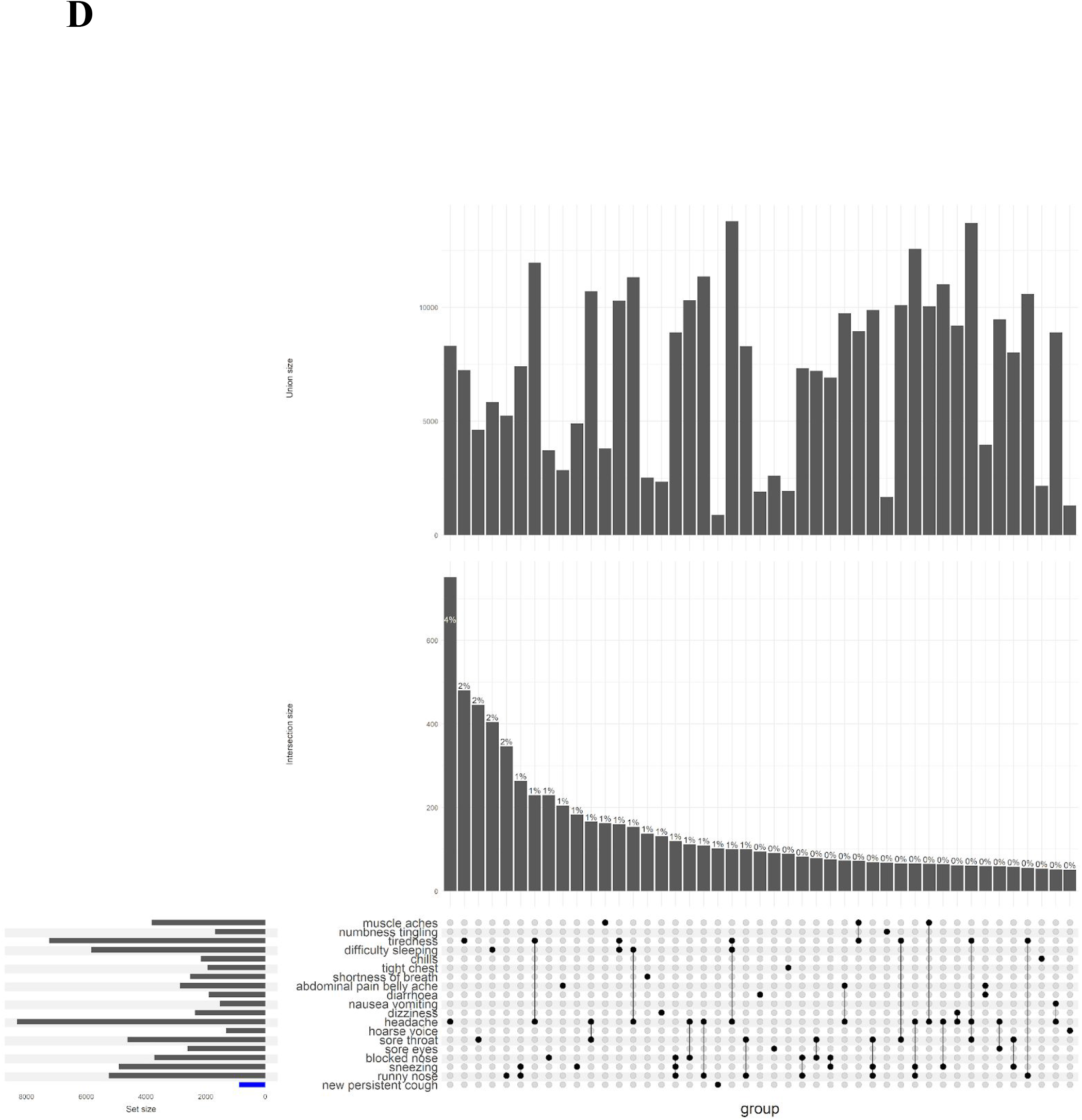
Upset plots showing frequency of (co-)occurrence of the 26 surveyed COVID-19 symptoms (or any combination thereof), ordered by frequency, among SARS-CoV-2 PCR positive (A) and negative participants (B) in rounds 2–7, and among PCR positive (C) and negative participants (D) in round 8. For readability, only (sets of) symptoms with >3 observations among rounds 2–7 positives and >200 observations among rounds 2–7 negatives are shown (and >3 and >50 in round 8, respectively). Union barplots (top panels) show the non-exclusive count of participants with that symptom profile – i.e. the symptoms in the set, plus any number of other symptoms, if applicable. Intersection barplots (middle panels) show the exclusive count of participants with that symptom profile – i.e. the symptoms in the set and *no other* symptoms.

**Supplementary Figure 3.**
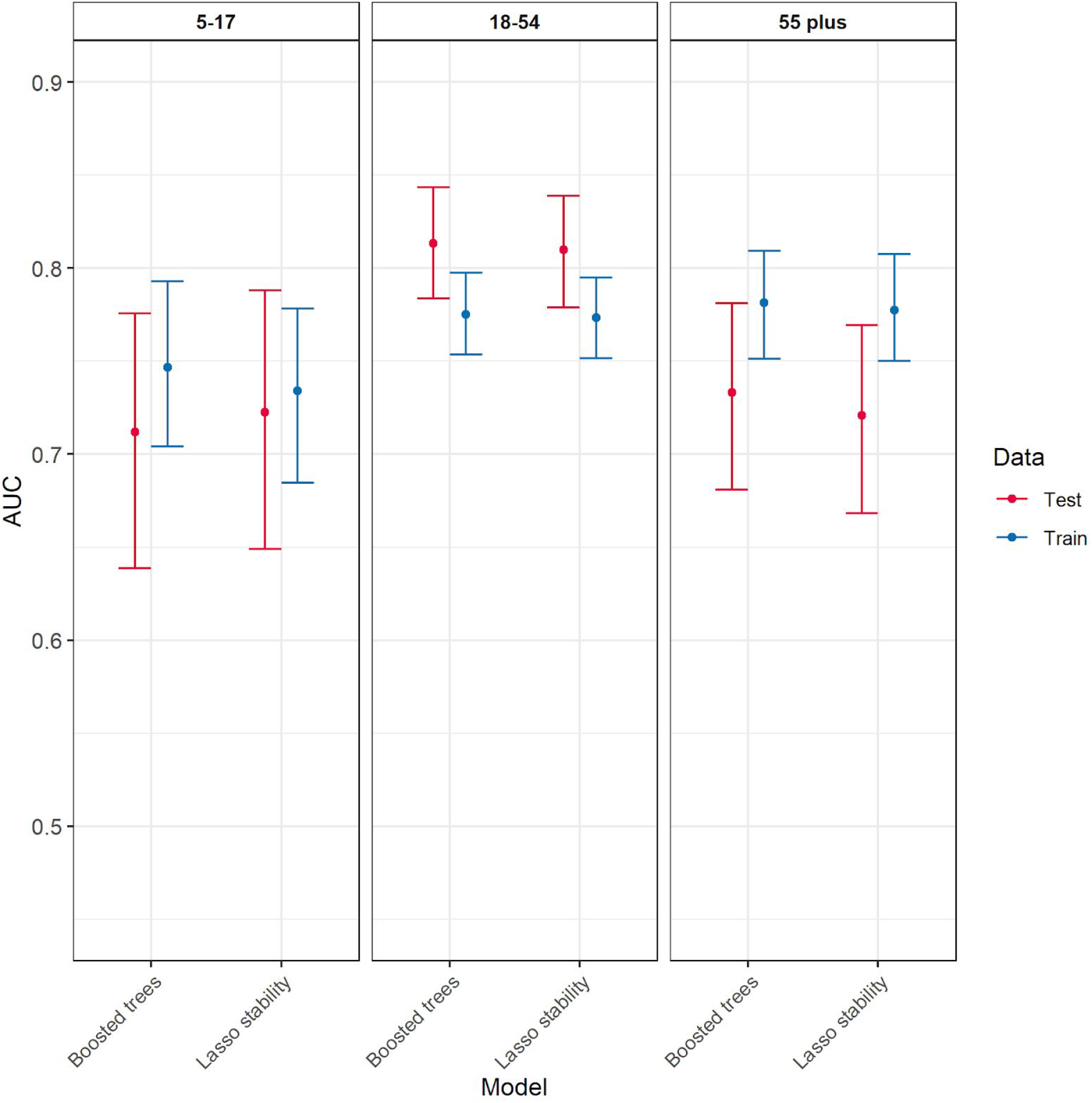
Comparison of model performance based on area under the receiver operating characteristic curve (AUC) using gradient-boosted forests (xgboost) versus our stability selection logistic LASSO approach. Results are presented for age-stratified training sets (70% of round 2 to 7 data, in blue) and evaluated on test sets (remaining 30% of round 2 to 7 data, in red).

**Supplementary Figure 4.**
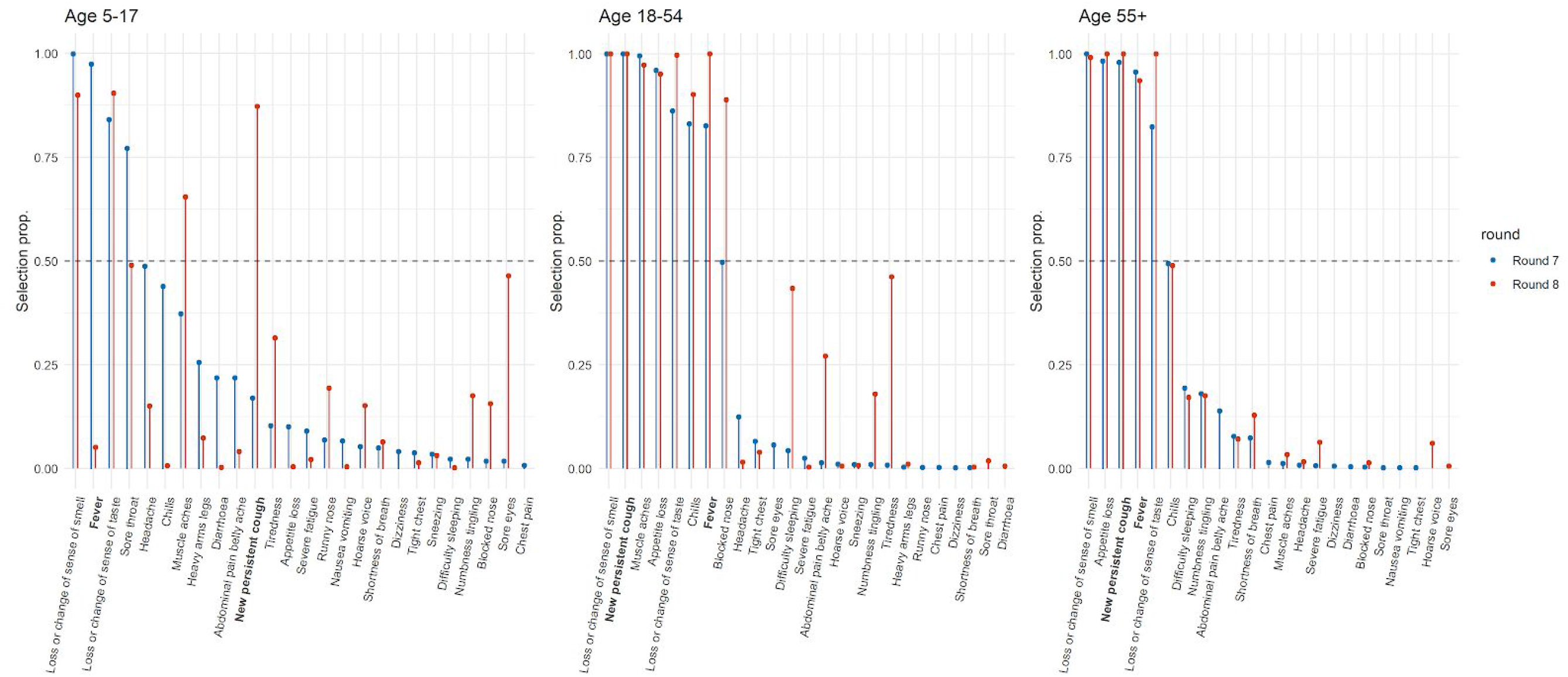
Results of the LASSO stability selection modelling in round 8 (in red) compared with round 7 (in blue) for the 5–17, 18–54 and 55+ year olds. Selection proportion for a given symptom is the number of times it was included in a calibrated logistic LASSO model (across 1,000 investigated 50% subsamples).

